# Aerosol emission rates from playing wind instruments – Implications for COVID-19 transmission during music performance

**DOI:** 10.1101/2021.12.08.21267466

**Authors:** C. Firle, A. Steinmetz, O. Stier, D. Stengel, A. Ekkernkamp

## Abstract

**Background:** The pandemic of COVID-19 led to exceeding restrictions especially in public life and music business. Airborne transmission of SARS-CoV-2 demands for risk assessment also in wind playing situations. Previous studies focused on short-range transmission, whereas long-range transmission has not been assessed so far.

**Methods and findings:** We measured resulting aerosol concentrations in a hermetically closed cabin of 20 m^3^ in an operating theatre from 20 minutes standardized wind instrument playing (19 flute, 11 oboe, 1 clarinet, 1 trumpet players). Based on the data, we calculated total aerosol emission rates showing uniform distribution for both instrument groups (flute, oboe). Aerosol emission from wind instruments playing ranged from 7 ± 327 particles/second (P/s) up to 2583 ± 236 P/s, average rate ± standard deviation. The analysis of the aerosol particle size distribution showed that about 70 − 80% of emitted particles had a size ≤ 0.4 µm and thus being alveolar. Masking the bell with a surgical mask did not reduce aerosol emission. Aerosol emission rates were higher from wind instruments playing than from speaking and breathing. Differences between instrumental groups could not be found, but high interindividual variance as expressed by uniform distribution of aerosol emission rates.

**Conclusions:** Our findings indicate that aerosol emission depends on physiological factors and playing techniques rather than on the type of instrument, in contrast to some previous studies. Based on our results, we present risk calculations for long-range transmission of COVID-19 for three typical woodwind playing situations.

## Introduction

The pandemic of COVID-19 has forced many countries to shut down major parts of their public life, including the music business. Virus-laden aerosols exhaled by infectious persons are a potential source of secondary SARS-CoV-2 infections ^1–4^. Resulting attack rates depend on a plethora of factors, the most significant of which are the aerosol emission rate of the primary case and the dilution factor ^5^. Singing indoors has early been found hazardous ^5–7^ and a correlation between voice activity level and aerosol emission rate is widely accepted ^8–10^. Since even silent breathing generates aerosols ^11^ it is necessary to examine to which extent wind instrument playing possibly increases particle emission over normal breathing, without vocalization. Since increasingly infectious virus variants have developed during the pandemic, this question remains relevant for the music scene.

Orchestras, ensembles, conservatories, and music schools have a demand for reliable data on aerosol emission from wind instrument playing ^12^. Protection from aerosol inhalation requires wearing FFP2 masks which is not feasible for the player of a wind instrument. Social distancing and face masks other than FFP2 protect susceptible persons from droplet transmission, but not from airborne transmission of SARS-CoV-2. Protective measures against aerosol transmission need to focus on ventilation and airing. Infection risk calculations involve the aerosol emission rate as primary model parameter.

Woodwind instrument playing by an infectious person imposes disease transmission risk on other persons in the same room at two different scales of distance: Short-range and long-range.

The primary aerosol cloud in the immediate vicinity of the player may carry pathogens in amounts sufficient to provoke quick infection of a susceptible person nearby. Within this nearfield of a spreader disease transmission occurs in a discontinuous manner difficult to predict ^13^ and dependent on both distance and contact time. Susceptible persons in this region are subject to what we call short-range exposure to infection risk.

While larger droplets (diameters > 50 µm) soon deposit due to gravitational settling or shrink to floatable aerosol particles ^14^, fine aerosol particles (with equilibrium diameters ≤ 5 µm) stay airborne and spread evenly in the surrounding air ^15,16^. In absence of technical measures, such as capture-type ventilation hoods, aerosols will accumulate in the room air at largely homogeneous concentrations. When infectious, they produce a base level of infection risk which increases over time and does not depend on a susceptible person’s position in the room ^1,5^. Susceptible persons in the room are subject to what we call long-range exposure to infection risk ^1,5^.

Previous studies of aerosol emission during woodwind playing gained important insight on the structure and extension of the nearfield cloud and helped to specify distancing rules for avoidance of short-range exposure. The flow structures in the vicinity of woodwind players have been investigated to characterize the shape, extensions and particle size composition of the primary exhalation cloud ^17–20^. The local air concentration of aerosol has been found to decay to background level on a sub-meter length scale which indicates efficient aerosol dilution in the ambient air ^19^. The plume generated by playing the clarinet has been found to be highly directional, to have high velocity, and to disperse quickly ^21^. No air motion could be measured in 2 m distance in front of woodwinds which was the largest reach of aerosol clouds observed ^17,18^. A reduction of aerosol emission by 50% (for particles in the size range 0.5 – 14 µm) to 79% (0.3 – 5 µm) when using a mask around the bell has been found^19,22^.

He et al. have assessed aerosol emission using aerodynamic particle sizers (APS) directly under the outlet of wind instruments to characterize the aerosol generation of different woodwind and brass instruments ^23^. Their results showed that playing wind instruments, in general, generates a higher number of aerosol particles than breathing and speaking. The amount of aerosol generated was dependent on various parameters, such as instrument type, dynamics, articulation and breathing techniques ^23^. This is consistent with the findings of Asadi et al. who investigated various speech components and demonstrated that aerosol production increases at loud speech or specific consonants ^24^. On the contrary, the results of McCarthy et al. suggest that playing wind instruments emits less aerosol than speaking and singing ^25^. Their study used aerodynamic particle sizers (APS) with funnels, like in previous studies ^10^. The categorization of wind instruments by aerosol emission as proposed by He et al. is not supported by McCarthy et al. ^25^. They discuss that the aerosol background concentration has a major impact on aerosol measurements and recommend clean room conditions.

None of the above-named studies focusing on short-range transmission allows to infer long-range transmission risk. Local aerosol concentration measurements cannot capture integral flow rates unless the total flow through a closed surface around the emission source is known. Local collection of emitted aerosol particles using under-pressure funnels ^22^ misses aerosol emanation elsewhere, without a chance to estimate the magnitude of the fugitive emission. This is less critical when measuring speaking or singing ^10^ but introduces systematic errors for instruments with keyholes, particularly the flute. Therefore, McCarthy et al. used two funnels when measuring aerosol emission during flute playing ^25^. Advanced collectors have been used by Stockman et al. to reduce the leakage errors during clarinet playing ^21^.

The observed quick dilution of aerosols entails concentration gradients which limit the accuracy of concentration measurements by adding an accidental dependence on the sampling position. Aerosol mass concentrations, as reported by Eiche ^26^, are affected by such local gradients and, additionally, by the nonstationary water content of the particles measured. Measurements of mass concentration depend on the particle size distribution and require calibration of the aerosol spectrometer for the specific kind of aerosol (solid versus liquid) ^27^.

The initial spontaneous motion of emitted aerosol particles is directed upwards, due to thermal convection, but when they reach the temperature of the surrounding air they mix uniformly with the latter. Eventually, aerosol particles spread all over the premises, following global air flow. The concentrations reached determine inhalation doses of potentially infectious aerosols.

For long-range transmission risk assessment, knowledge of the total emission rate of airborne aerosol particles is required. Measurement of the spatially uniform aerosol concentration, developing after sufficient mixing time inside a hermetic probe volume, allows calculation of the aerosol emission rate. The entire aerosol released by the proband is diluted in the known amount of air inside the probe cabin and the total emission rate is proportional to the temporal increase of the aerosol concentration. The aerosol emission rate is a property of the proband and not subject to spatial fluctuation. Leakage errors, as by airflow through keyholes, do not occur during such a measurement.

Dependency on the setting, in particular the ceiling height, has been proposed as possible cause of discrepancy between the results of Abraham et al. ^19^ and Kähler and Hain ^28^. The latter study found a larger extension of the local flow zone. Low headroom accelerates the lateral spreading of aerosols and the formation of uniform concentrations in a probe volume. Accordingly, our setup uses a hermetic probe cabin with low ceiling for accelerated formation of uniform concentrations. Moreover, our probands deliver realistic music performance rather than playing selected single notes ^25^.

The aim of this study is to measure aerosol generation under standardized conditions during the playing of different wind instrument groups. The primary research goal is to measure the total emission rate of airborne aerosol particles (< 20 µm diameter *in statu nascendi*) while playing wind instruments, including interindividual variation. A secondary question is whether super-spreaders could be defined based on their aerosol emission rate. Lastly, ways to reduce aerosol emission were examined (outlet cover). We use the measured emission rates to predict the probability of COVID-19 transmission in three typical woodwind playing situations.

## Methods

### Study design and participants

An exploratory, cross-sectional design was used to measure aerosol emission during wind instrument playing. A convenience sample of volunteer flute, oboe, clarinet, and trumpet players was sought from professional orchestras and students at music conservatories. To qualify for the study musicians had to be at least 18 years of age and to have had professional instrumental education (university degree or similar). Exclusion criteria included acute or severe chronic respiratory diseases and symptoms of a SARS-CoV-2 infection. Recruitment of the participants was accomplished by an informing homepage and an online registration form and was conducted between the end of August and end of October 2020. The measurements took place during October and the first week of November 2020.

All measurements were conducted with the approval of the relevant ethics committee of the University Medical Center of Greifswald (reference number BB 131/20) in accordance with the declaration of Helsinki. Every participant gave informed written consent. The study was registered retrospectively on the German Clinical Trials Register (www.drks.de) as No. DRKS00023336.

### Experimental setup

The measurements took place in an operating theatre of the outpatient surgery centre of Unfallkrankenhaus Berlin, Germany, to create quasi clean room conditions. Room temperature and relative humidity in the theatre were constant, controlled by air conditioning. Room air was primarily filtered by carbon filters and by F7 and F9 bag filters meeting the criteria of EN 779.

A measuring tent, the probe cabin, was set up in the operating theatre. It consisted of a 20 m^3^ cuboid constructed of aluminium bars and impermeable foil (see Figure 1) to prevent air exchange between the inner of the cuboid and the outside during measurements. Between measurements the stale air inside the cuboid was exchanged by opening the probe cabin at both ends. A fan and an air purifier with HEPA filter (high-efficiency particulate air H13, CARD 210 m^3^/h) accelerated the air exchange (see Figure 1). All participants were clothed in a surgical gown and hood and locked in after entry from the side of the probe cabin. During measurements, the upper part of the body was inside the probe cabin, tightly enfolded around the waistline by the sealing foil.

**Figure 1.**
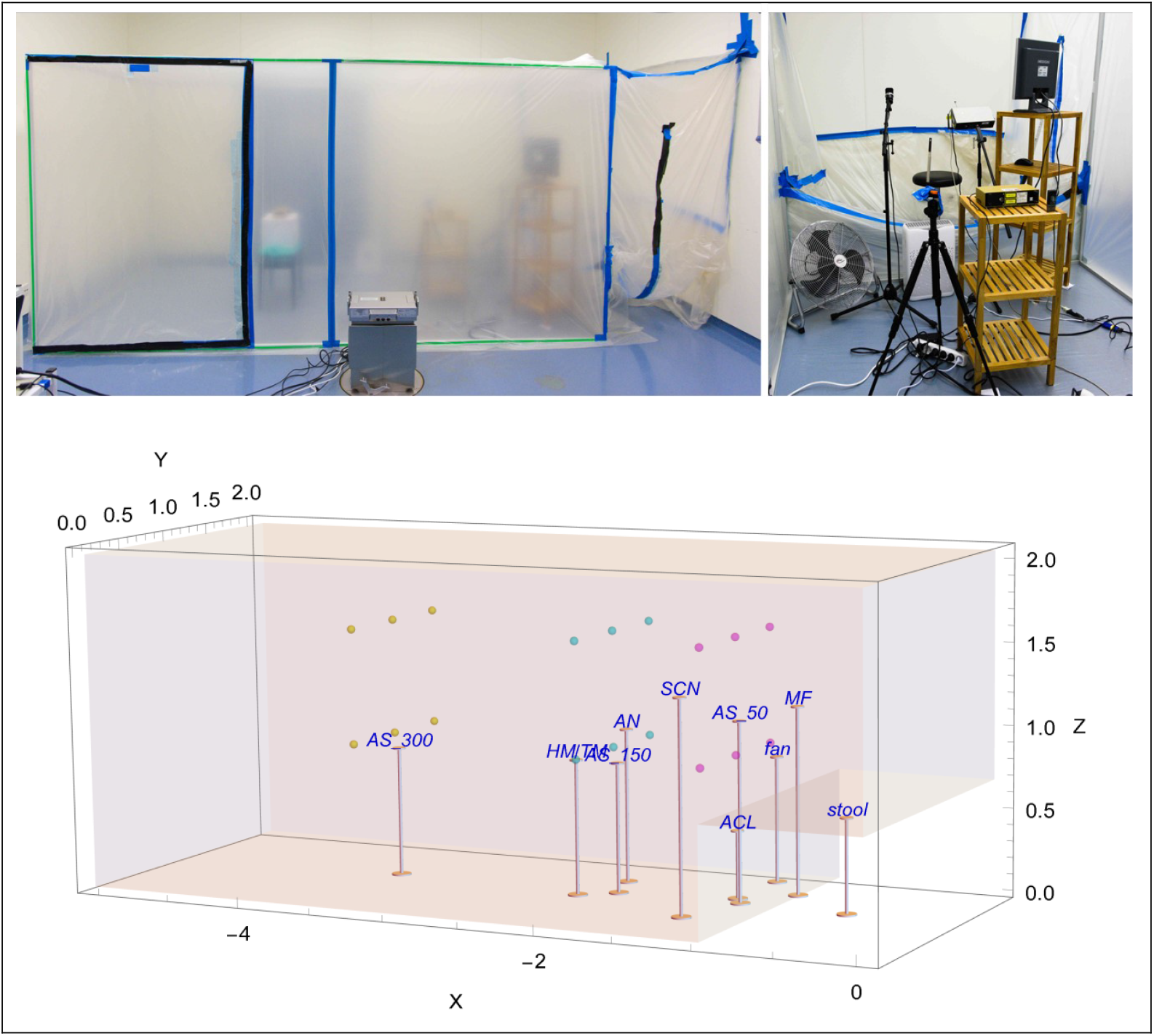
Photographs (top) and 3D-figure (bottom) of the experimental setup. Black tape on the probe cabin’s foil indicates the left and right opening slots allowing air exchange between successive measurements. Air flow measurement points are marked as coloured dots. Average room temperature was 24.0 ± 0.3 °C; average relative humidity was 42 ± 5% for flutists and 40 ± 6% for oboists. Air velocity never exceeded 0.01 m/sec at the anemometer position. AS = aerosol spectrometer, AN = anemometer, HM/TM = hygrometer/thermometer, MF = microphone, SCN = screen, ACL = air cleaner, scales in m.

Spatially resolved measurement of air speed inside the closed cuboid with one participant sitting inside indicated < 0.01 m/sec at all positions (measured by a testo 405 i anemometer, see Figure 1). Three aerosol spectrometers (Grimm Aerosol Spectrometers 1.109 and 11-D) were placed in 50 cm (11-D), 150 cm (1.109) and 300 cm (1.109) distance from the participant. A computer monitor was placed to the participants’ left sight showing scores or text. To record the sound as audio-track a microphone was placed on the right. The thermo-hygrometer (Voltcraft DL-220THP) and the anemometer were placed next to the aerosol spectrometer positioned at 150 cm distance, see Figure 1. Device specifications are reported in Appendix 2.

Aerosol concentrations were measured by the three aerosol spectrometers in the unit particles/litre, categorized by size into 31 bins ranging from >0.25 µm to >35 µm, at collection intervals of 6 seconds. Relative humidity (RH, unit: %) and air velocity (unit: m/s) were measured at 150 cm distance from the participant. Every 2 seconds air speed was recorded, and every 60 seconds temperature and humidity. All devices recorded continuously during a whole session, the start and end times were logged by a html-java-script.

### Tasks

Flute and oboe players were asked to play Mozart’s Oboe Concerto in C major K. 314 or Mozart’s second Flute Concerto in D major K. 314, respectively (same concerto, originally arranged for flute by Mozart himself). Clarinet and trumpet players played a piece of their own choice ^29^. Each recording lasted 20 minutes. Subsequently, participants were asked to read a text (the beginning of Hermann Hesse’s “Der Steppenwolf”, in German) for 20 minutes, followed by 20 minutes of quiet breathing.

### Data evaluation

Measurement of total particle emission rates during flute and oboe performance is complicated by the non-uniform flow of exhaled air which simultaneously emanates from different openings of the instrument and the mouth of the player. The total flow is, thus, difficult to measure whereas local particle measurements near the instrument would sample an unknown fraction of the total emission. Therefore, we measured the spatially uniform aerosol concentration developing after sufficient mixing time inside a hermetic probe volume of 20 m^3^. The entire aerosol amount released by the proband is diluted in the known amount of air inside the probe cabin. The total emission rate is proportional to the increase of the average aerosol concentration inside the cabin.

Since the aerosol spectrometers 1.109 and 11-D use slightly different particle size bins, the latter were harmonized by bin fusion forming the new size bins >29.9, >25.1, >12.8, >6.6, >3.0, >1.6, >0.8, >0.4, >0.25 µm appearing in the figures below. Particles with diameters up to 6.6 µm are further combined to the size bin “aerosol” whereas larger particles form the bin “droplets”. Details of the emission rate calculation are reported in Appendix 1 and in the document “data processing.pdf” in our repository ^29^.

The aerosol emission rates determine inhalation doses of potentially infectious aerosol particles and allow long-range transmission risk calculations (Appendix 3).

Data evaluation and statistics were performed with Wolfram Mathematica 12.2 (www.wolfram.com/mathematica). Data sets, supplementary data and figures are available in our repository ^29^.

## Results

Descriptive and demographic data of participants are reported in Table 1.

**Table 1.** Descriptive and demographic data of musicians. Mean (standard deviation) and number of cases are reported. py = pack years

|  |  | Flute player | Oboe player | Clarinet<br>player | Trumpet<br>player |
| --- | --- | --- | --- | --- | --- |
| n |  | 19 | 11 | 1 | 1 |
| gender identity | female : male | 16 : 3 | 5 : 6 | 0 : 1 | 0 : 1 |
| Age [years, 5-year<br>range] | mean | 47 (15) | 51 (12) | 26-30 | 26-30 |
| height of<br>participants [cm] | female | 166 (6) | 168 (11) |  |  |
|  | male | 182 (3) | 188 (9) | 174 | 182 |
| weight of<br>participants [kg] | female | 65 (12) | 66 (9) |  |  |
|  | male | 81 (6) | 84 (11) | 75 | 90 |
| respiratory disease | none | 18 | 11 | 1 | 1 |
|  | allergy-induced<br>bronchial asthma | 1 |  |  |  |
| nicotine<br>consumption | none | 17 | 7 |  | 1 |
|  | previously | 1 (3 py) | 2 (1 py, 3 py) |  |  |
|  | currently | 1 (12 py) | 2 (7 py, 35<br>py) | 1 (2 py) |  |
| years of experience |  | 36 (14) | 38 (11) | 20 | 22 |
| beard | no | 18 | 8 |  |  |
|  | three-day beard | 1 | 3 |  |  |
|  | full beard |  |  | 1 | 1 |

### Total aerosol emission rates and particle size distribution

Aerosol emission rates of a total of 32 musicians playing four different wind instruments covered the range from 7 ± 327 particles/second (P/s) to 2583 ± 236 P/s, average rate ± standard deviation (see Figure 2a for details). The aerosol emission rates from playing clarinet and trumpet lay within the lower range of the uniform distributed aerosol emission rates from playing flute and oboe.

**Figure 2.**
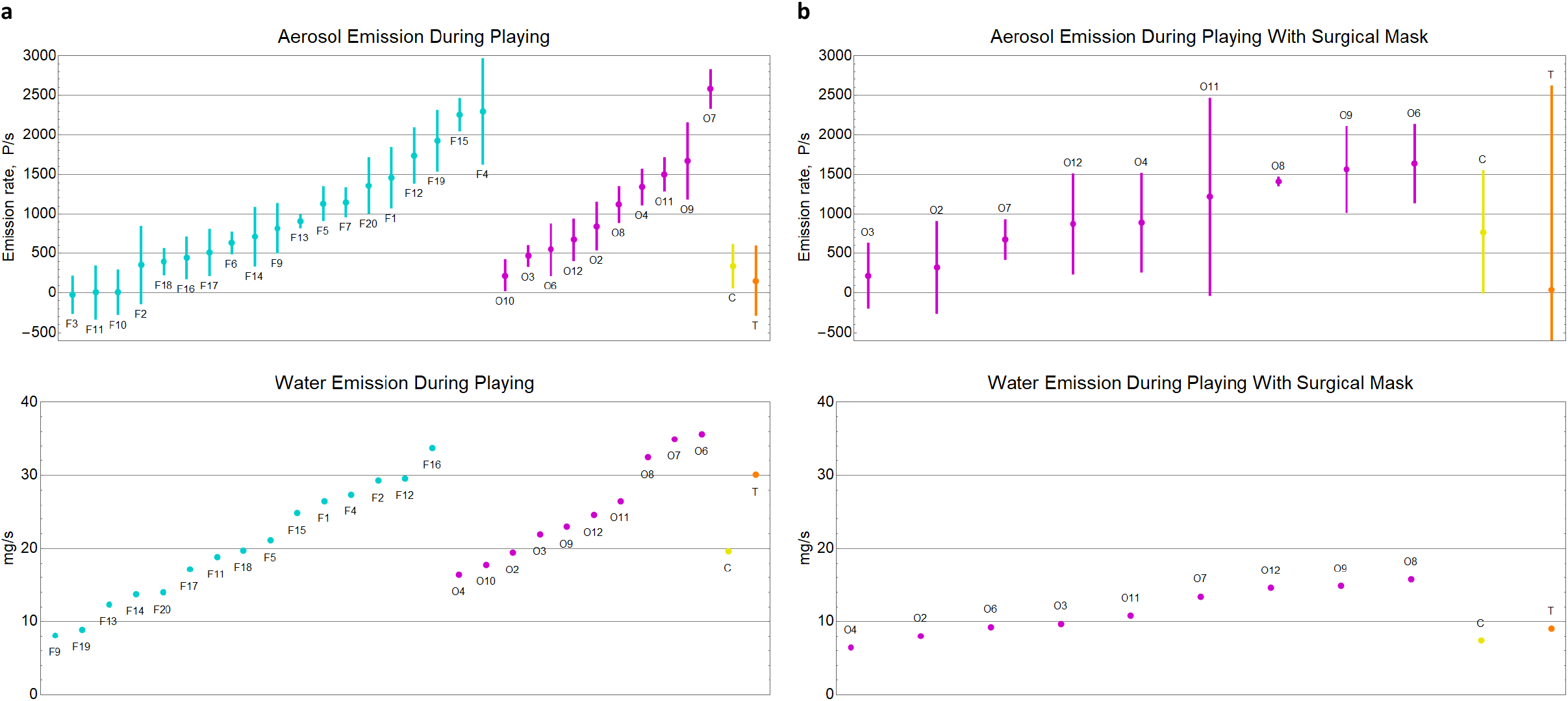
Distribution of aerosol and water emission rates during wind instrument playing without (a) and with (b) a surgical mask around the bell. F = Flute, O = Oboe, C = Clarinet, T = Trumpet. Numbers indicate participant’s ID. Wind instrument playing lasted 20 minutes; oboe, trumpet, and clarinet playing with a surgical mask lasted 10 minutes. Emission rates are reported as particles/second (P/s), water emission rates as mg/s. Error bars indicate standard deviations according to bootstrap estimation.

Analysis of the aerosol particle size distribution (see the histogram in Figure 3) shows that about 70 − 80% of the emitted particles had a size ≤ 0.4 µm. Particles larger than 6.6 µm (droplets) were rarely detected. Differences between the size distributions of aerosols from different instruments cannot be seen. Since particles smaller than 1 µm penetrate deeply into the respiratory tract, down to the alveolar parenchyma, this aspect is of particular importance with regard to the risk assessment of wind instrument playing.

**Figure 3.**
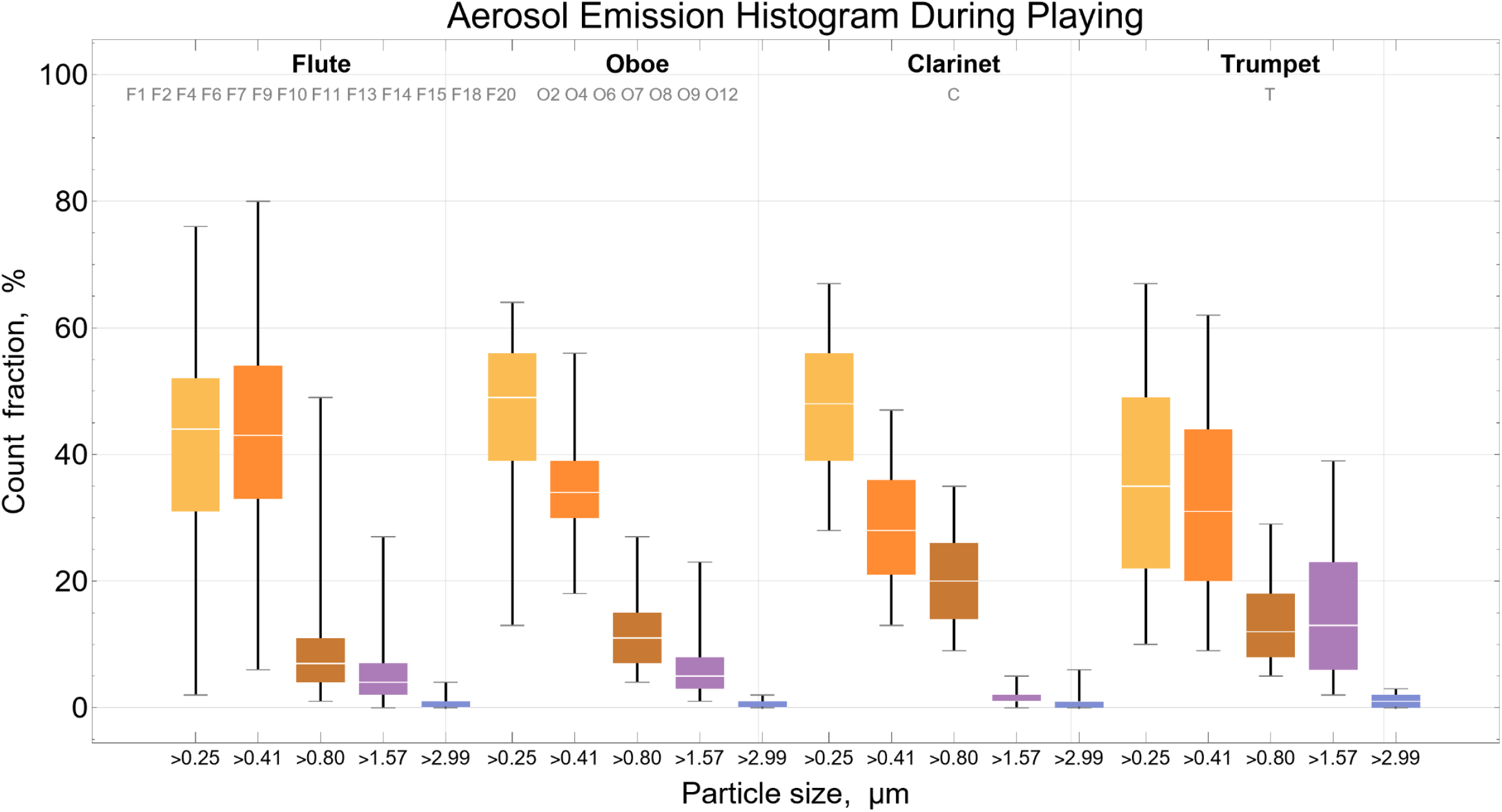
Size distribution of aerosol from playing wind instruments. White split indicates median, whiskers indicate lower and upper fence, box indicates 25% and 75% quantiles.

### Aerosol emission rates of playing wind instruments compared to speaking and breathing

Aerosol emission rates from music playing were found to be higher than those from speaking and breathing (see Figure 4). Interestingly, the sole exception (oboist no. 12) shows similar emission rates for both music playing and speaking (673 ± 253 P/s oboe playing, 800 ± 128 P/s speaking). Aerosol emission rates from speaking varied up to 800 ± 128 P/s and from breathing up to 566 ± 352 P/s. Individual aerosol emission rates from speaking were consistently higher than from breathing. The distributions of emission rates from speaking and breathing are uniform.

**Figure 4.**
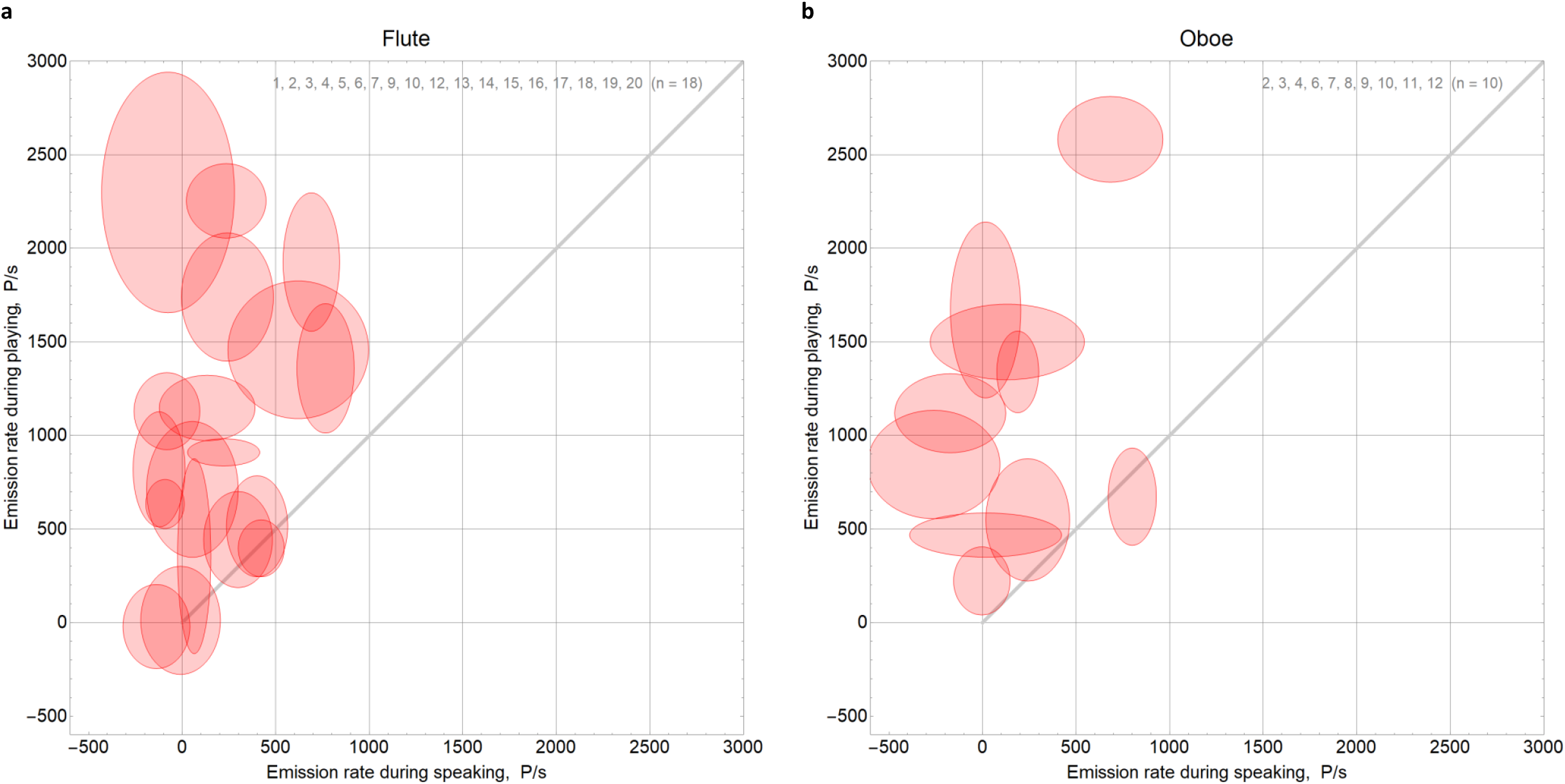
Aerosol emission rates (unit: particles/s) are displayed for the tasks wind instrument playing (a) flute, (b) oboe, and speaking. The centre point of the ellipse indicates the emission rate from instrument playing on the ordinate and from speaking on the abscissa. The semiaxes of the ellipse indicate the standard deviations for each task, according to bootstrap estimation. The grey line indicates equal emission rates at both tasks. Ellipses above that line indicate higher emission rates from wind instrument playing than from speaking.

### Aerosol protection by using a bell mask

In attempt to reduce aerosol emission, the oboists, the clarinettist, and the trumpeter played with a surgical mask over the bell for 10 minutes. Due to the shorter measurement time, the relative standard deviations of aerosol emission rates are higher. We find no difference between the aerosol emission rates from playing with and without mask. Figure 2b shows a uniform distribution of the aerosol emission rates from playing with a surgical mask around the bell.

Figure 5 compares the aerosol emission rates from playing with and without mask. The semiaxes of the ellipses indicate the standard deviations. Despite large standard deviations, individual aerosol emission rates are consistent between both tasks (playing with and without mask), except for one outlier showing the expected reduction of emission by the mask (oboist no. 7).

**Figure 5.**
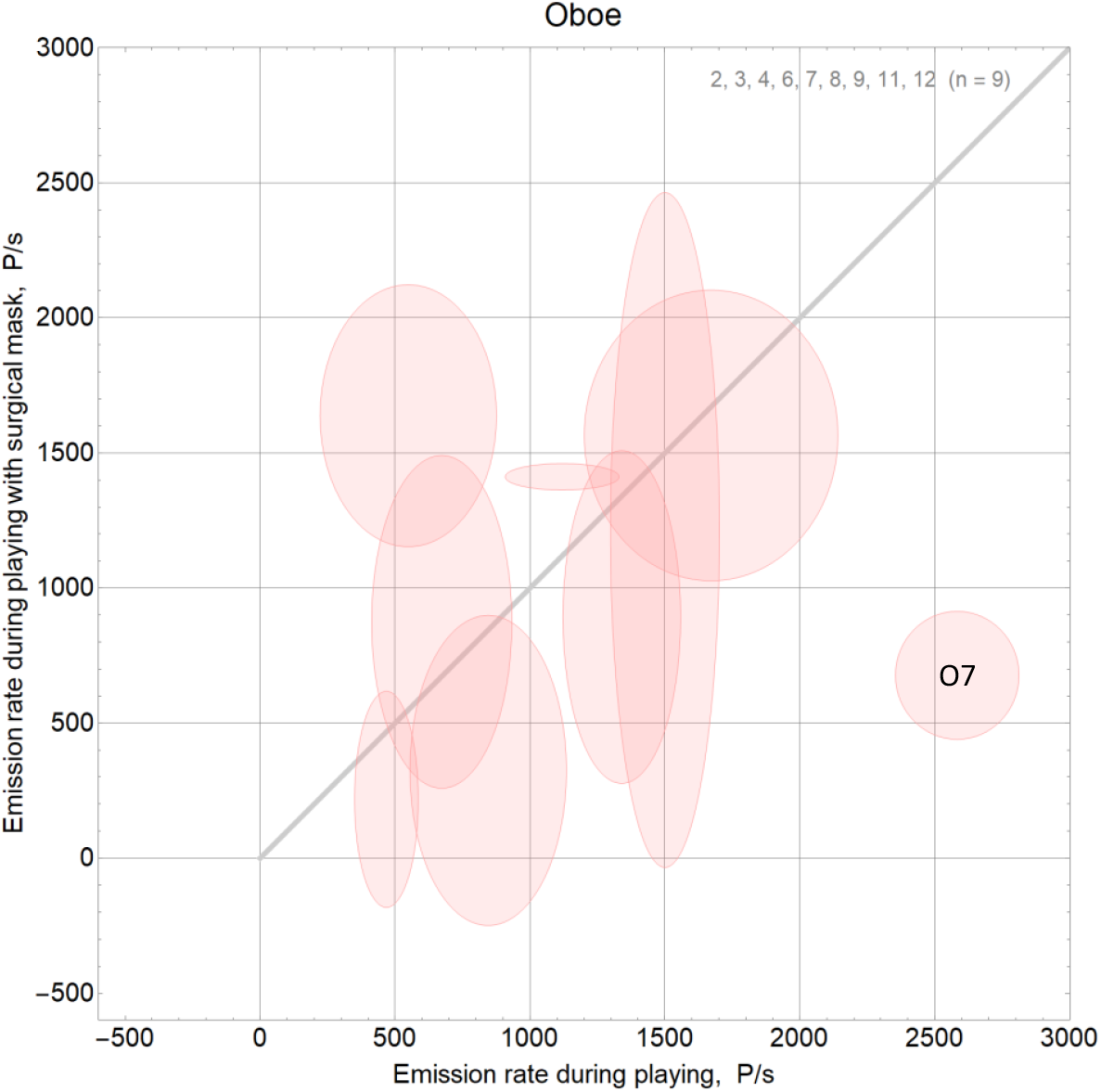
Aerosol emission rates (unit: particles/s) for oboe playing with and without a surgical mask around the bell. Only proband O7 shows essential reduction in aerosol emission when playing with a surgical mask.

### Water emission

Complementary to the total aerosol emission rates, we calculated the total water emission rates from the continuous increase of relative humidity during task performance. The water emission rates show a uniform distribution, like the aerosol emission rates (see Figure 2b). To elucidate the correlation to aerosol emission we calculated the ratio and find that 1 mg emitted water corresponds to ≤ 100 emitted aerosol particles. The ratio was smaller for the task speaking, with two exceptions considered dry high emitters (see Figure 6).

**Figure 6.**
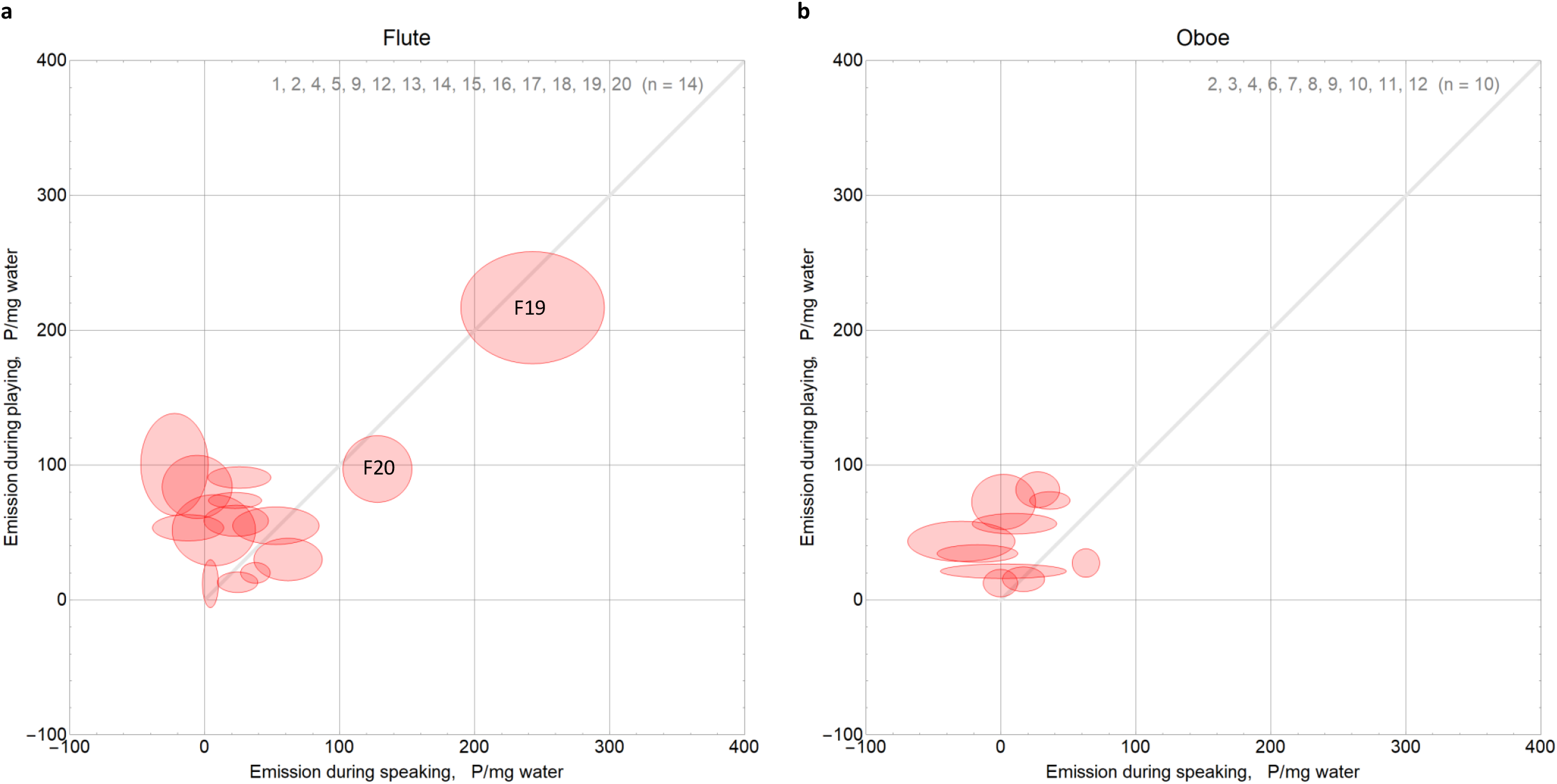
Particle emission per mg water emission while playing (a) flute, (b) oboe, and speaking. Particles per water ratios are higher from wind instrument playing than from speaking in both groups. F19 and F20 are dry high emitters.

Since the humidification of exhaled air takes place in the upper respiratory tract we assume that water emission rates provide important information about the respiratory volume and its circulation in airways. We plotted water emission rates during instrument playing versus speaking and found a high correlation with factor 2.2 for all wind instruments, trumpet and clarinet included (see Figure 7 and correlation plots in our repository ^29^). This indicates that water emission universally increased by factor two from speaking to wind instrument playing. The same holds for the comparison of instrument playing to breathing, yielding a factor four. If water emission correlates with respiratory volume, the relation of pulmonary ventilation rates during breathing, speaking, and wind instruments playing appears to be very similar for all participants.

**Figure 7.**
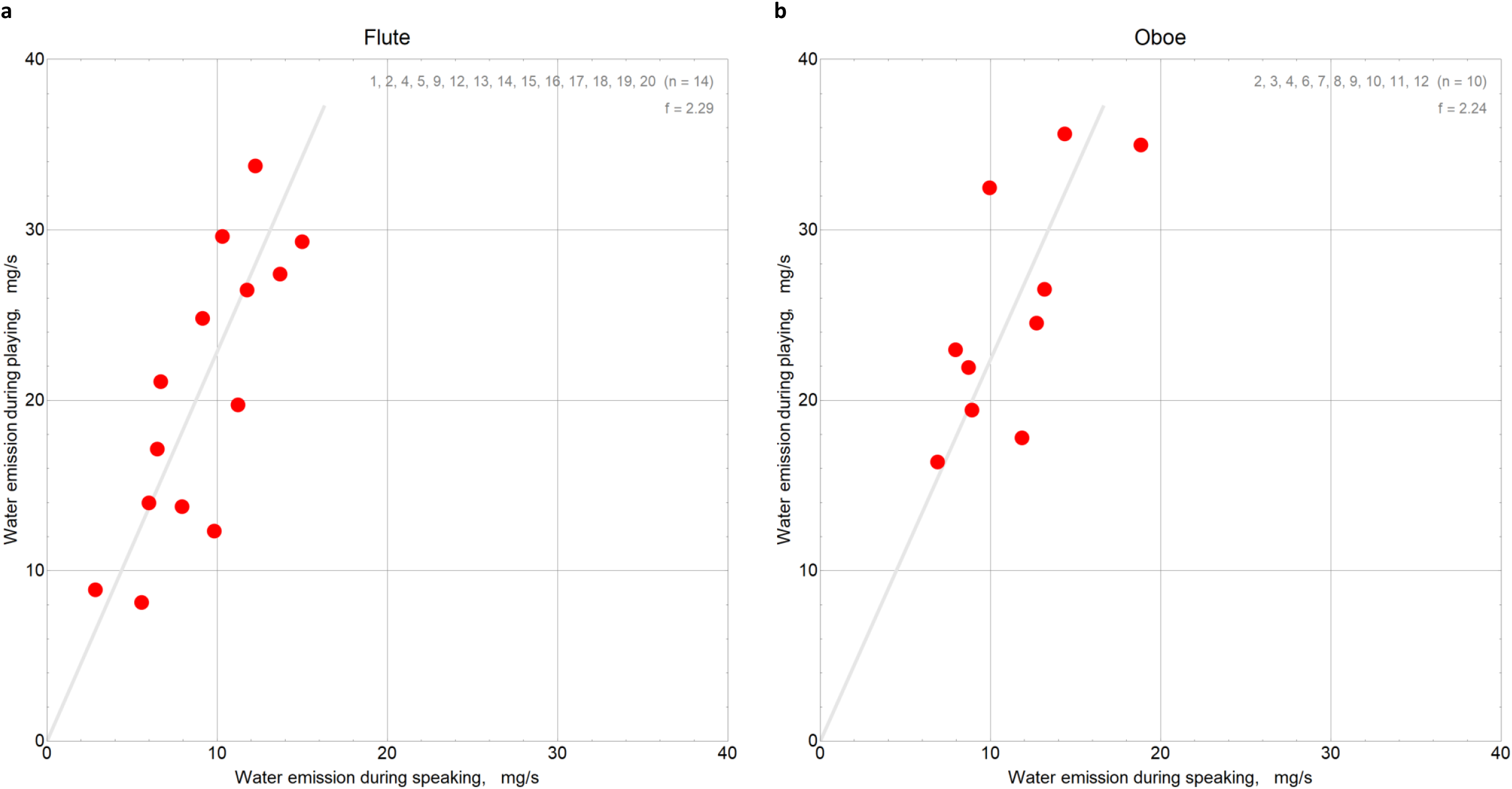
Water emission rates (unit: mg/s) while playing (a) flute, (b) oboe, and speaking. Regression lines have virtual identical slopes of 2.2 for both groups, i.e., water emission doubles from speaking to wind instrument playing.

## Discussion

To our knowledge, this study was the first to examine more than two representatives of the same instrument, but a greater and thus more representative number of individuals for two wind instruments (oboe, flute). Moreover, the study design using a hermetically closed probe cabin and standardized playing condition allowed, for the first time, measurement of total aerosol emission rates. Other than in previous studies ^23,25^ the different musicians performed the same repertoire piece of music including a variety of dynamics and articulation techniques. Our experiment resembles a realistic performance situation with respect to both the scores and playing time (20 min).

Our results show that wind instrument playing generates higher aerosol emissions than speaking or calm breathing. We observe total emission rates in the range previously reported for singing, exceeding 1000 particles per second ^30^. This does not support a recent study reporting higher local aerosol concentrations from speaking and breathing than from wind instrument playing ^25^. Our probands played a whole Mozart Concerto rather than single notes ^25^ and we attribute our different findings to this difference between the two experiments: Performance of a long piece activates the typical breathing techniques to greater extent than sustaining tones for 20 s. Our results agree with He et al. who found that playing wind instruments in general generates more aerosol than breathing and speaking, whereby the emission rate is dependent on parameters, such as dynamics, articulation, and breathing techniques ^23^. This is in line with findings of Asadi et al. investigating various speech components and demonstrating that aerosol production increases with speech loudness or use of specific consonants ^24^.

The large number of probands playing oboe and flute in our study demonstrated major individual variability within the two groups. Emission rates show uniform distribution within similar ranges for the two instruments. Unlike previous studies ^19,20,23^, no clear allocation of emission rates to the instrument type is possible. We conclude that individual factors dominate the variability of aerosol emission rather than the type of instrument. Outliers from the uniform distribution that might be interpreted as super-spreaders could not be found, other than in a previous study that detected high aerosol emitting probands during speaking ^8^.

In search for individual factors influencing the aerosol emission we found that emission rates do not correlate with body height or weight ^29^. Hence, we assume that the breathing technique is probably the reason for individual variability of aerosol emission.

The humidification of exhaled air takes place in the upper respiratory tract ^31,32^ whereas aerosol formation is thought to originate deeper in the respiratory tract ^11^. Since the humidified air is saturated with water even at high flow rates ^33^, water emission likely correlates with pulmonary ventilation rate. Our results reveal higher aerosol particles per water ratios for wind instrument playing than for speaking. Assuming wind instrument playing requires higher pulmonary ventilation rate than speaking, our results are consistent with an increased air exchange in the respiratory tract during wind instrument playing. The required pulmonary volume apparently depends on individual factors, such as vital capacity or breathing technique, which explains the high variability of aerosol emission within the two instrument groups. We found noteworthy correlations between the water emission rates from wind instrument playing, speaking, and breathing indicating that the respiratory volume needed for the respective task might increase uniformly for all the different individuals.

Regarding the particle size distribution, most of the particles are < 1 µm in diameter, as previously found for breathing and speaking probands ^30^. Particles > 6.6 µm in diameter were rarely recorded, thus being negligible for long-range, airborne disease transmission, in agreement with results of McCarthy et al. ^25^. SARS-CoV-2 virus has a diameter of 0.13 µm ^34,35^ and saturation of the smallest aerosol particles has been shown ^36,37^. This imposes great risk for long-range transmission since particles < 2 µm reach alveolar parenchyma.

Like other researchers before, we tried to reduce the aerosol emission by masking measures. We masked the bell with a surgical mask on the oboe, clarinet, and trumpet. Except for one oboist, all participants produced similar aerosol emission rates as without mask. A previously described reduction of 50% to 79% ^19,22^ was not observed at the measurement distances used in our study. We assume that aerosol emanated through keyholes and embouchure.

### Risk assessment of typical woodwind playing situations

Short-range exposure is difficult to model, but easy to mitigate (by social distancing following recommendations, e.g., in Gantner et al. and Hedworth et al. ^18,38^). The opposite applies for long-range exposure, in practice. The obvious countermeasures against aerosol transmission are ample fresh air and the wearing of FFP2 masks. However, the efficiency depends strongly on the specific setting. The sole simple rule available is the recommendation to do outdoor whatever can be done outdoor. For indoor occupation, COVID-19 transmission risk can be calculated as described in Reichert et al. ^5^ and implemented online for free use: https://hri-pira.github.io^39^.

We apply the framework outlined in Appendix 3 to assess the criticality of a few typical situations of playing woodwinds. It is assumed that appropriate social distancing excludes short-range exposure so that the infection risk entirely results from long-range exposure. As mentioned before, the hazard in a particular scenario depends on both the individual aerosol emission rate *q* and the infectiousness of the instrument player. In a given real situation the disease transmission probability may be a factor 10 less than stated below, or even negligible, since we subsume the worst case.

For our calculations we assume the maximal infectiousness (*Z*_50_ = 833 particles, for the Delta variant) and an aerosol emission rate of *q* = 2500 particles per second while playing, to examine whether *the setting* is safe or not. This question remains important even when an antigen test has been carried out before playing since asymptomatic spreaders may pass at significant rates reported with a sensitivity of 58% to 95% ^40^. A safe setting provides the necessary, second line of defense. Vaccination is neglected in the following, thus assuming susceptibility for infection.

### Lesson at the music school

The teacher and an infectious student have a 60 min lesson in a 200 m^3^ classroom. The student listens 50%, plays 40%, and talks 10% of the time (average aerosol emission rate *q* = 4·10^6^ /h). Neither wears a mask and the windows remain closed. The resulting long-range infection probability for the susceptible teacher is *p* = 96%. To reduce *p* to 10% by ventilation only, unrealistic 80 air changes per hour (ACH) sustained were necessary. If, instead, the teacher wears a tight FFP2 mask with a filter efficiency of 95% (*ϑ* = 0.05) ^41,42^ then *p* = 15%. They may open the door and windows widely for 10 min after half an hour to clear the air from aerosols. Then, *p* = 79% without wearing a mask, or *p* = 7% wearing a mask. To conclude, acceptable safety levels can be reached even at worst-case conditions by

i. limiting the duration to one hour,
ii. wearing FFP2 mask whenever suitable, and
iii. obligatory, thorough airing around half time.

Infection probability with mask is expected around 14% when the space volume of the room is half as large (100 m^3^).

### Recital

An infectious soloist plays a one-hour program (net playing time) accompanied by two musicians in a 2000 m^3^ ballroom. The audience leaves the room after 90 minutes, including the encores and applause. Automatic ventilation exhausts air through the ceiling at 2 ACH (4000 m^3^/h) with fresh air streaming inward near the floor. The CO_2_ level stays below 1000 ppm (good air quality) for audiences up to 100 persons. The average aerosol emission rate is *q* = 6·10^6^ /h. The long-range infection risk for susceptible persons is *p* = 3% if they wear FFP2 masks and *p* = 45% otherwise. Given that social distancing prevents accommodation of more than 50 spectators in the ballroom, one secondary infection case is expected when FFP2 masks are worn throughout. The reproductive number in this setting is *R* ≈ 1.

### Symphonic performance

A woodwind player in an orchestra is infectious. They play symphonic literature, i.e., the duty cycle of woodwinds is average. We assume 30 min net playing time evenly distributed over 90 min concert duration, resulting in an average aerosol emission rate *q* = 3·10^6^ /h. The concert hall has a space volume of 20000 m^3^. Automatic ventilation exhausts air above stage and auditorium at 2.5 ACH. The long-range infection risk for susceptible persons is *p* = 0.14% when they wear FFP2 masks, and *p* = 3% otherwise, owing to the large air space and fresh air supply.

The basic assumption of full mixing, or perfect aerosol dilution, is questionable in the latter example. Concert stages may have air exhaustion ducts which remove part of the air on stage from the hall before it mixes into the air surrounding the audience. In case of the opposite flow direction, fresh air streaming down from the ceiling, problems may arise, such as local stagnation or recirculation regions with elevated aerosol concentrations ^38^. Generally, large premises require consideration of actual flows and should not be assessed using the well-mixed room air assumption. Our example cannot be generalized to other concert halls based only on their size and total air supply.

## Conclusion

Wind instrument playing, like singing, should be considered more dangerous than speaking or breathing. High interindividual variance of emission rates indicates that physiological factors and playing techniques shape the extent of aerosol formation, rather than the type of instrument. To investigate influencing factors of individual aerosol emission further studies are needed. Our results pave the way to absolute risk calculations for oboe and flute playing in typical situations.

## Data Availability

All data produced in the present work are contained in the manuscript and in our repository at https://github.com/Carl-Firle/Aerosol_Study_Wind_Instruments.git

## Acknowledgments

We wish to thank the Unfallkrankenhaus Berlin and especially Dr. Hajo Schmidt-Traub and Jörg Karst with his team at the Outpatient Surgery Centre for their help in conducting the study.

We wish to thank Dr. Udo Jäckel and Daphne Bäger with the team at the Bundesanstalt für Arbeitsschutz und Arbeitsmedizin Berlin for their sustained assistance in measurement procedure and the noteworthy development of the study design.

We thank Dr. Chan Yong Schüle for his consultancy in air flow physics and the development of the experimental setup.

Finally, we thank the Deutsche Orchester Vereinigung for their help in the recruitment process.

## Financial support

This study was supported by a research grant of the German Association of Music Physiology and Music Medicine, Germany.

## Conflict of Interests / Competing interests

Oliver Stier is named as inventor on a patent application filed recently regarding the determination of potentially infectious aerosol air concentrations.

All authors declare no competing interests.

## Author contributions

Conceptualization, CF and AS; methodology, CF, AS, DS, creation of models OS; validation, CF and OS; investigation, CF; formal analysis, OS; software, CF and OS; resources, AS, AE; writing—original draft preparation, CF, AS, OS; writing—review and editing, CF, OS, AS; visualization, OS; supervision AS, AE; project administration, CF, AS; funding acquisition, CF and AS. All authors have read and agreed to the published version of the manuscript.

## Appendix 1

### Measured quantity

Prerequisite for correct measurement of total emission is that the quantity of aerosol does not change between origin and detection, i.e., along the path from the proband’s respiratory tract to the spectrometer. Once formed, exhaled aerosol particles undergo significant changes of their water content by competing processes of hygroscopic growth and desiccation ^14^. It takes in the order of a second for an airborne particle to attain a stable equilibrium diameter. Any earlier measurement of particle sizes or masses would yield nonstationary, irreproducible quantities while later sampling would map the equilibrium state rather than the original one.

In contrast, the number of particles is preserved from formation to detection, thus being the more reliable measure for amounts of exhaled aerosols. The spectrometers we used return both mass and number distributions. We consider the latter since mass values in most cases require model-based rescaling to the original particle size which would introduce speculation to the measurement result. The size distributions we obtained have undergone an unknown, but continuous, transformation and depend on the relative humidity inside the probe cabin. Most droplets larger than 30 µm in diameter have got lost due to gravitational settling. The counts of smaller, airborne particles are well comparable to other measurements at the same relative humidity.

### Measurement accuracy

The signal-to-noise ratio (SNR) of particle counting measurements is intrinsically limited by random count fluctuations resulting from finite volume sampling. The number *n* of aerosol particles of same kind inside the spectrometer sampling volume is a random draw from a Poisson distribution whose expectation value is proportional to the particle air concentration. In our setup, the sampling volume for measurement of particles larger than 2.5 µm was 100 ml air and for those smaller than 2.5 µm (vast majority of particles) 20 ml air, at 1.2 l/min sampling flow rate and 6 s duration per measurement cycle. Detection of *n* particles smaller than 2.5 µm indicates an air concentration of 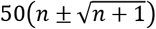 particles/litre (P/l), with the square root expression being the standard deviation of estimation uncertainty ^43^. *n* particles larger than 2.5 µm indicate a concentration of 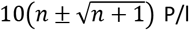. The calculation applies to every size bin separately (kind of particle). For example, a concentration of 150 P/l for diameters 0.7 − 0.8 µm is measured with an uncertainty of ± 100 P/l since *n* = 3. More precise values are obtained by averaging dozens of single values. This entails long measurement times and leads to the chosen performance duration of 20 min per task.

During this period each of the three spectrometers measures 204 count values in time intervals of 6 s. These are averaged in groups of 34 subsequent values to obtain six new data points equidistant in time. The 34 single values are measured within a period of 200 s which is the time scale for uniform distribution of aerosol particles inside the probe cabin. By assumption of well mixed room air simultaneous particle counts at the three spectrometers are draws from the same Poisson distribution representing the global particle concentration at that time. Thus 3·34 = 102 original count values coalesce to one air concentration value whose noise is Gaussian and reduced by factor 10, compared to single count values.

A straight line representing the expected linear increase of particle air concentration over time is fitted to the six consolidated data points. Its slope, multiplied by the cabin volume, is the point estimate of the time-averaged total particle emission rate of the proband. The emission rate error bar is calculated using the bootstrap method ^44^ and given as the standard deviation of the marginal distribution of the emission rate.

**Appendix 2**

### Device specifications

*Dust-Decoder 11-D* and the *Portable Aerosol Spectrometer Model 1*.*109* measure with a total inlet flow of 1.2 l/min (20 ml/s). A diode laser counts particle numbers during the 6 second interval (120 ml air sample) using light scattering. Particle concentrations up to 2.000.000/liter can be detected. The smallest concentration resolution is 1 particle / 20 ml for particles from 0.25 - 2.5 μm, and 1 particle / 100 ml for particles from 2.5 – 32 µm.

The thermo-hygrometer *Voltcraft DL-220THP* has a resolution of temperature of 0.1 °C, of relative humidity 0.1%, and of air pressure 1 hPa. The measuring range of temperature is −30 to 60 °C, of relative humidity 0 to 100%, of air pressure 300 to 1200 hPa. Accuracy of temperature is 0.5 °C, of relative humidity 3.5%, and of air pressure 2 hPa.

The anemometer *testo 405 i* senses air flow with a resolution of 0.01 m/sec. The measuring range is 0 to 30 m/sec, the accuracy is ± 0.1 m/sec + 5% of the mean for velocities between 0 and 2 m/sec and ± 0.3 m/sec + 5% of the mean for velocities of 2 to 15 m/sec.

The microphone used was *TLM 102 bk* produced by Neumann. It was linked to the Zoom recorder H4n Pro. Output was saved as mp3-file with a bit rate of 128 kbps.

**Appendix 3**

### Calculation of transmission risk

The exponential dose-response model relates the probability *p* of infection to the intake dose of pathogens, *Z*, by

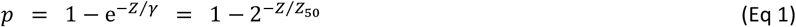

where *γ* is a disease-specific, characteristic dose referred to as ‘quantum of infection’ ^45^ and *Z*_50_ = *γ* ln 2. The common unit of *Z, Z*_50_, and *γ* is free to choose; it could be, for instance, RNA copies, plaque-forming units, or aerosol particles. *Z*_50_ is neither a threshold dose, nor a tolerance dose, but determines how fast infection risk statistically increases at increasing inhalation dose *Z. Z*_50_ quantifies the average susceptibility of non-vaccinated individuals to infection by the respective virus strain. If the pathogen concentration of the inhaled air is constant over time, Eq. (1) can be written as the Wells– Riley equation ^46^.

Recently, *Z*_50_ has been estimated for SARS-CoV-2 as number of (partly) virus-laden aerosol particles whose equilibrium diameters are in the range 0.3 µm to 5 µm ^5^, the same size range as in our present work. This makes it possible to use the aerosol emission rates determined here for calculation of absolute COVID-19 transmission probabilities in situations typical for woodwind playing: A lesson at the music school, a recital, and a symphonic performance in a concert hall. These scenarios involve non-stationary accumulation of aerosols in the air which is not described by the Wells–Riley equation.

For the SARS-CoV-2 lineage B.1.1.7 (Alpha) *Z*_50_ = 1166 (median, 95% CI: 700 − 1942) aerosol particles have been proposed as likely lower bound for the characteristic dose of infection ^5^. The lineage (Delta) has a factor 1.4 − 1.6 higher transmissibility than Alpha ^47–49^. This factor is composite of a higher infectiousness (expressed by *Z*_50_) and a longer duration of infectiousness (resulting in more opportunities for secondary infections). We assume an increase of infectiousness over the Alpha variant by factor 1.4 and apply the consideration of Reichert et al. to derive *Z*_50_ = 833 aerosol particles with equilibrium diameters 0.3 µm − 5 µm for the Delta variant ^5^.

The infectious power of a given number *Z* of aerosol particles depends on the number of active virus contained. For SARS-CoV-2, highest infectiousness has been shown between two days before and the day of symptom onset ^50–53^. The virion concentration varies by one or two orders of magnitude, depending on the individual and the infection stadium ^54,55^.

Accordingly, transmission risk calculations based on the above *Z*_50_ value apply to the worst case that the woodwind player performs during the phase of maximal infectiousness. Few days earlier, or a week later, the total viral load of aerosol particles released could be a factor 10 to 100 smaller. Hence, our calculations only indicate how dangerous instrumental play could be in the extreme case. They are apt to assess the criticality of environments and the efficacy of safety measures, whereas they cannot specifically predict case numbers of secondary infection in the context of music performances.

The inhalation dose *Z* in Eq. (1) is the number of (partially) infectious aerosol particles inhaled by a susceptible person during the period of exposure. It is calculated assuming instant, perfect mixing between aerosol and room air as outlined in Reichert et al. ^5^. *Z* may be scaled by the inward transmissivity of a face mask worn which reduces infection probability by the factor *ϑ* = 1 – (filter efficiency). When no mask is used, *ϑ* = 1, and *Z* remains unaltered.

Influencing factors of the long-range infection risk *p* are the use of FFP2 masks for self-protection (*ϑ*) and the

1. total emission rate of infectious aerosol particles, *q*
2. exposure duration
3. room space volume
4. ventilation or airing.

In our study we quantify the first factor. To observe the scatter between individuals we measured *q* for 19 flute and 11 oboe players performing the same task under identical conditions. The second to fourth factor depend on the scenario, as outlined in the discussion chapter.

## Notes

### Competing Interest Statement

The authors have declared no competing interest.

### Funding Statement

This study was supported by a research grant of the German Association of Music Physiology and Music Medicine, Germany.
No author and no institution involved in the project received payment or services from a third party at any time.

### Author Declarations

The relevant ethics committee of the University Medical Center of Greifswald (reference number BB 131/20) gave ethical approval for this work in accordance with the declaration of Helsinki.

